# Diagnosing Rejection Collapse via Uncertainty Decomposition

**DOI:** 10.64898/2026.07.24.26358775

**Authors:** Walter Endrizzi, Flavio Ragni, Stefano Bovo, Monica Moroni, Giuseppe Jurman, Venet Osmani

## Abstract

Standard uncertainty-informed rejection can unexpectedly trigger severe performance collapse, exposing localized vulnerabilities that common machine learning metrics typically do not show. We systematically diagnose this failure dynamic using Levodopa-Induced Dyskinesia prediction in Parkinson’s Disease as a proof-of-concept. By training a heterogeneous ML ensemble, decomposing Aleatoric and Epistemic uncertainty and applying unsupervised subgroup discovery, we isolated the precise drivers of these atypical errors. Stratified error analysis revealed two divergent predictive regimes previously hidden by a global evaluation. While the models successfully extracted a predictive signal for one subgroup, the baseline features of a second subgroup lacked discriminative capacity, resulting in a high rate of confident misclassifications. Operating entirely below rejection thresholds, this single subgroup flatlined predictive metrics, driving the collapse of the global rejection curve. Ultimately, we demonstrate that atypical rejection failures stem from subgroup-specific data ambiguity rather than algorithmic deficiencies, making localized uncertainty-aware evaluation a critical methodological requirement prior to real-world deployment.

## 1 Introduction

Despite algorithmic advancements, the transition of predictive models into clinical practice is hindered by the opaque nature of standard ML evaluations. Prognostic models are predominantly assessed using global performance metrics, which aggregate results across heterogeneous patients. Not only do these aggregated scores obscure localized, subgroup-specific vulnerabilities, but standard metrics also fail to quantify predictive confidence. Consequently, it becomes impossible to determine whether errors stem from algorithmic deficiencies, insufficient training data, or irreducible ambiguity inherent to the clinical variables themselves. Beyond merely identifying these errors, safely deploying clinical ML requires models to actively utilize their own uncertainty. In standard practice, this is achieved through performance-rejection mechanisms, which operate on the assumption that discarding a model’s most uncertain predictions will monotonically increase performance on the remaining cohort. However, this assumption no longer holds if a model produces highly confident misclassifications. Disentangling the sources of predictive uncertainty is therefore essential, shifting the focus from simply evaluating model architecture to interrogating the true prognostic utility of the clinical data itself.

To demonstrate the practical impact of disentangling this clinical uncertainty, we focus on the prognostic challenges of Parkinson’s Disease (PD). Levodopa remains the most effective pharmacological treatment for managing motor complications in PD. However, its long-term administration is frequently complicated by the development of Levodopa-Induced Dyskinesia (LID), a debilitating condition characterized by involuntary movements that severely impact a patient’s quality of life. The ability to forecast an individual patient’s risk of developing LID using only baseline clinical assessments would fundamentally transform PD management, enabling clinicians to tailor initial therapeutic strategies and delay LID onset [1]. Consequently, machine learning (ML) prognostic models have increasingly been applied to clinical datasets to identify early predictive markers of LID [2].

In this study, we propose an uncertainty-informed framework as our primary contribution, using the forecasting of LID from real-world, first-visit clinical data as a proof-of-concept to rigorously assess predictive limits. By training a heterogeneous ML ensemble and decomposing predictive uncertainty using Information Theory, we expose a severe, atypical performance collapse during rejection analysis. Leveraging unsupervised subgroup discovery, we demonstrate that this predictive failure is not a systemic algorithmic flaw, but is strictly localized to a specific subgroup of early-stage patients.

## 2 Data and Methods

### 2.1 Data and clinical task

The dataset comprised baseline evaluations of 247 PD patients recruited across two Italian clinical centers, incorporating demographic, clinical, and motor-symptom assessments [3]. The binary classification task was formulated to predict the onset of Levodopa-Induced Dyskinesia (LID) within a three-year window. To prevent the models from learning a trivial classification shortcut, the target outcome at baseline (LID status at baseline) was explicitly removed from the input feature space. Furthermore, to simulate a true prognostic scenario, all final performance metrics and uncertainty estimations were computed exclusively on the clinically relevant subgroup of patients who did not present with LID at baseline (*N* = 212).

### 2.2 Base model training and nested Cross-Validation

Model training and hyperparameter optimization were executed using a Randomized Nested Grid Search Cross-Validation (CV) pipeline. The entire workflow was iterated 30 times. In each outer iteration, the data was randomly partitioned into an 80% training set and a 20% hold-out test set stratified by the target outcome and baseline motor-complication status. Within each of the 30 training loops, a repeated (5 repetitions) 3-fold CV approach was employed to optimize hyperparameters over a randomized search space of 20 configurations. Five base ML algorithms were optimized: Random Forest (RF), ExtraTrees Classifier (ETC), Extreme Gradient Boosting (XGB), Logistic Regression (LR), and Support Vector Classifier (SVC). Additionally, two intra-iteration ensemble techniques were added to this pool: a Voting classifier aggregating the predictions from RF, SVC, XGB, and LR, and a Stacking classifier utilizing XGB, RF, and SVC predictions as input features for a final LR meta-model. This yielded a total of 7 distinct classifiers trained per iteration. The optimal base model configuration selected in each iteration underwent probability calibration prior to evaluation. Platt Scaling was applied using an internal 5-fold CV scheme on the training data. Calibration quality was visually verified using calibration curves to confirm the alignment between predicted probabilities and observed empirical frequencies [4, 5].

### 2.3 Global ensembling and uncertainty quantification

Following probability calibration, a global heterogeneous ensemble was constructed post-hoc to leverage the diverse hypothesis spaces of the algorithms. This global ensemble aggregated the calibrated predictive probabilities from all 7 models across all 30 iterations. By pooling these test-set predictions, we treated the *M* total predictions for any given patient as the ensemble’s predictive distribution [6]. Predictive uncertainty was decomposed into its aleatoric and epistemic components based on the established predictive distribution. We adapted an information-theoretic framework to quantify uncertainty in generalized ML spaces and tree-based ensembles [6, 7]. For a given patient, let *p*_*m*_ denote the calibrated predictive probability of LID onset generated by the *m*-th constituent model of the global ensemble, where *m* ∈ {1, …, *M*}. For example, in our 210-model ensemble (7 classifiers × 30 iterations), *p*_1_ = 0.72 could represent the calibrated probability of LID onset predicted by Random Forest during its first iteration. Total uncertainty (H_*total*_) is calculated as the predictive Shannon entropy of the global ensemble’s consensus probability 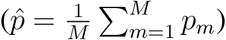, capturing all sources of predictive doubt:

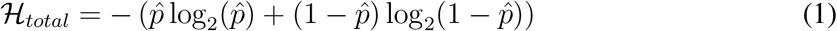

The Aleatoric uncertainty (*H*_*aleatoric*_) is formulated as the expected entropy across the ensemble members [6]. High values identify specific regions within the feature space where the available baseline variables possess insufficient discriminative power to reliably separate the classes:

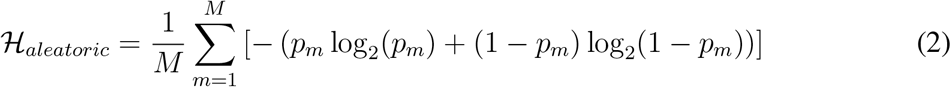

Epistemic uncertainty (*H*_*epistemic*_) is derived as the mutual information between the predictions and the model parameters, calculated as the difference between Total and Aleatoric uncertainty [7]. This captures uncertainty arising from finite training data or out-of-distribution inputs:

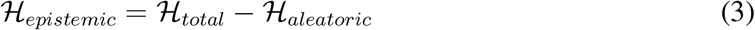

### 2.4 Uncertainty-informed rejection analysis

To evaluate whether the computed uncertainty estimates reliably map to classification performance, we conducted a rejection evaluation using Classification-Rejection Curves (CRCs). This analysis tests the hypothesis that higher predictive uncertainty correlates with a higher likelihood of misclassification. Patients who appeared at least once in the hold-out test sets were ranked according to their Total uncertainty. We iteratively removed the top *k*% of the most uncertain patients (from 0% to 80% rejection thresholds) and re-evaluated the global ensemble’s performance on the remaining subset. Performance at each threshold was quantified using the Matthews Correlation Coefficient (MCC) to account for any class imbalances induced by the rejection process.

### 2.5 Unsupervised subgroups discovery

To prevent data leakage, clustering was performed strictly on the imputed (mean imputation) and standardized baseline clinical features, remaining completely blind to the models’ risk predictions and true clinical outcomes. Dimensionality reduction was first applied using Principal Component Analysis (PCA), retaining exactly 9 principal components (eigenvalue *>* 1.0) according to the Kaiser Rule. A Gaussian Mixture Model (GMM) was then fitted to this reduced subspace to identify data-driven patient subgroups. The optimal cluster count (*k*) was determined by jointly evaluating the Bayesian Information Criterion (BIC) to penalize complexity and the Silhouette score to maximize inter-cluster separation.

### 2.6 Clinical profiling and rejection analysis

Once the unsupervised clusters were established, they were mapped back to their original clinical variables. A comparative clinical profiling was conducted using two-sided Mann-Whitney U tests to identify statistically significant baseline features defining each subgroup’s clinical signature. Finally, the uncertainty-informed rejection analysis was stratified by the discovered clusters: Subgroup-specific CRCs were computed across increasing rejection thresholds, tracking the evolution of predictive performance via the MCC, sensitivity, and specificity.

## 3 Results

### 3.1 Global predictive uncertainty and the rejection paradox

The global heterogeneous ensemble demonstrated that predictive uncertainty was driven almost entirely by data ambiguity rather than model ignorance. As illustrated in the uncertainty phase-plane (Figure 1A), Epistemic uncertainty remained negligible across the cohort (*<* 0.06), indicating robust consensus among the base classifiers across all iterations. In contrast, Aleatoric uncertainty spanned the full theoretical range up to 1.0. A vertical concentration of predictions near *H*_*aleatoric*_ ≈ 1.0 contained a highly intermixed population of actual positives and actual negatives, confirming that the baseline clinical features of these specific patients inherently lack the discriminative capacity to resolve the clinical outcome. Furthermore, the CRCs revealed an unexpected performance degradation (Figure 1B). Instead of increasing monotonically as highly uncertain predictions were discarded, the MCC dropped from 0.38 to negative values at rejection thresholds greater than 70%. This atypical dynamic indicates that patients who remained at high rejection rates were not simply reliable classifications, but rather confident misclassifications (i.e., instances where the ensemble was highly certain, yet entirely incorrect).

**Figure 1:**
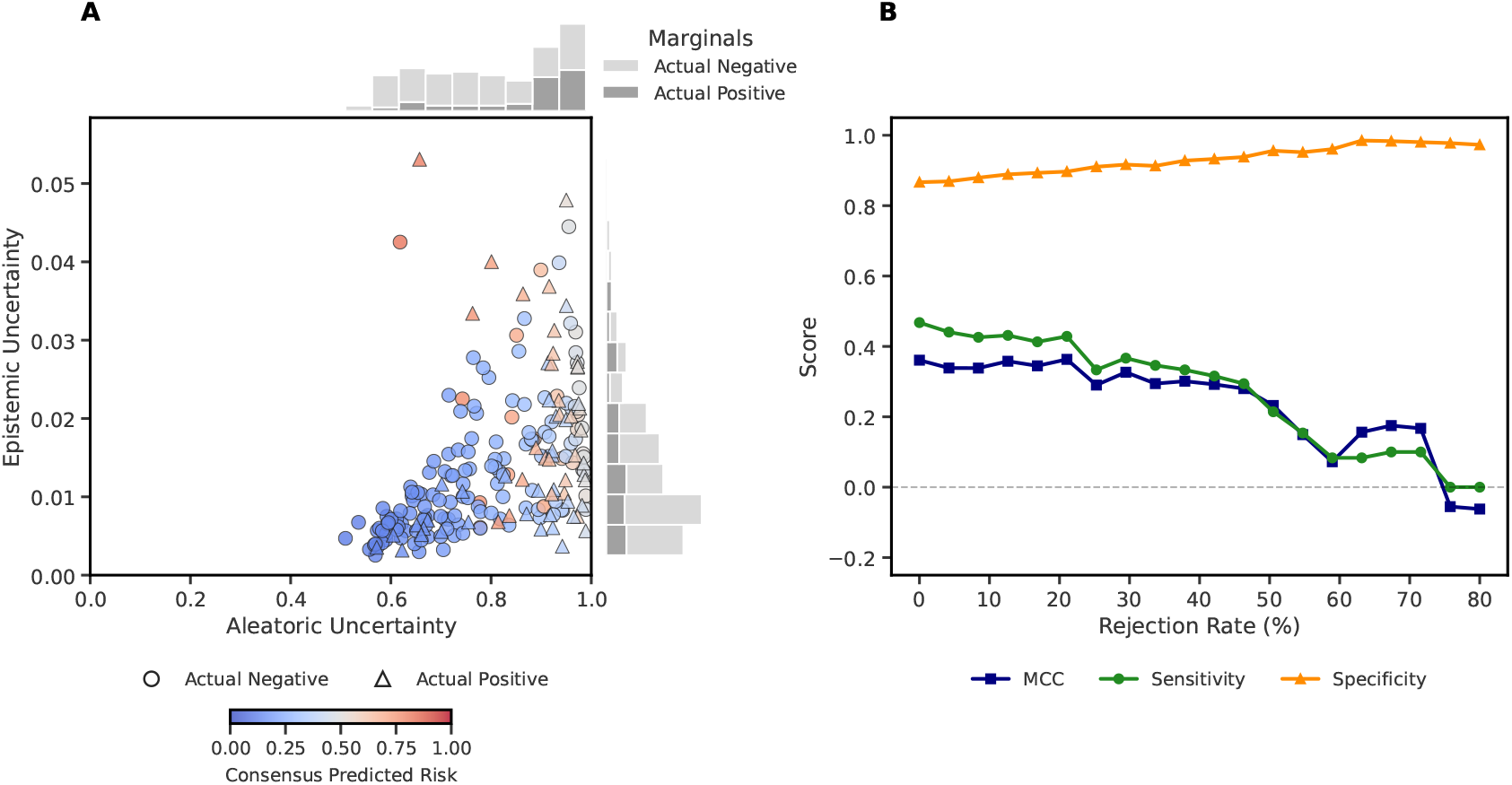
Global uncertainty landscape and atypical rejection dynamics. **(A)** The uncertainty phase-plane demonstrates that model ignorance (Epistemic uncertainty) is negligible across the cohort, whereas data ambiguity (Aleatoric uncertainty) spans the full theoretical range. The intermixed target classes at high aleatoric values highlight regions where baseline clinical features lack discriminative power. **(B)** The Classification-Rejection Curve for Total uncertainty. Discarding uncertain predictions counterintuitively degrades performance, causing the MCC to collapse from 0.38 to negative values at high rejection thresholds, indicating a subgroup of confident misclassifications.

### 3.2 Data-driven subgroup discovery

To determine if specific patient profiles were driving this atypical performance collapse, unsupervised clustering was applied exclusively to the baseline clinical features. Dimensionality reduction via PCA retained 9 components according to the Kaiser rule. Subsequent GMM, optimized via BIC and Silhouette scores, strictly identified two distinct patient subgroups (*k* = 2). Clinical profiling utilizing two-sided Mann-Whitney U tests mapped these mathematical clusters back to their defining clinical signatures. Subgroup A (*N* = 77) exhibited a significantly more advanced disease profile compared to the mild subgroup B (*N* = 135) across different key baseline features (all *p <* 0.001). Specifically, subgroup A presented with longer disease duration (1.03 vs. 0.24 years), higher Levodopa Equivalent Daily Dose (449.6 vs. 164.4 mg), more advanced Hoehn & Yahr stages (2.05 vs. 1.65), and a vastly higher baseline prevalence of motor fluctuations (38% vs. 2%) and freezing of gait (27% vs. 0%).

**Table 1:**
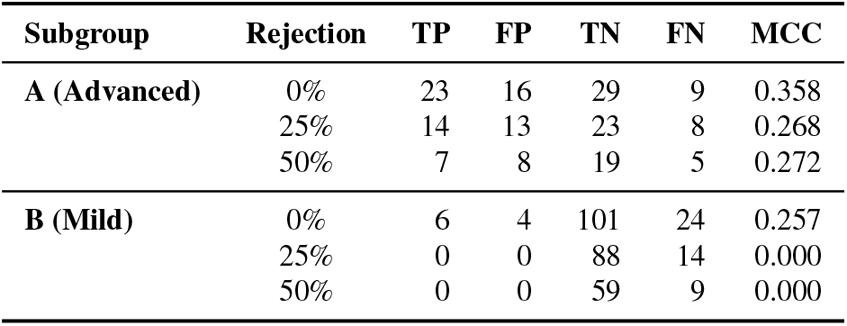
Confusion matrix evolution across rejection thresholds.

### 3.3 Stratified error analysis

Conducting a stratified error analysis on these discovered subgroups revealed the precise root cause of the global model collapse. As detailed in Table 3.3, tracking the localized evolution of the confusion matrices demonstrates that the dataset’s discriminative failure is not a uniform algorithmic deficiency, but a subgroup-specific limitation. For subgroup A (Figure 2A) the ensemble maintains its ability to identify both LID and non-LID even under extreme rejection. At a 50% rejection rate, the model successfully identifies 7 True Positives (TP) and 19 True Negatives (TN). While the MCC does not monotonically increase, the model avoids systemic failure, indicating that advanced baseline symptoms provide a limited but reliable prognostic signal that the algorithms can successfully exploit. In stark contrast, Subgroup B (Figure 2B) exhibited a severe confidence-error dynamic. The model’s behavior rapidly collapsed as uncertainty thresholds were applied. By the 20% rejection mark, the ensemble completely ceased predicting the positive class, causing the MCC to mathematically flatline to 0. The remaining confident pool consisted exclusively of 88 True Negatives and 14 False Negatives. This explicitly proves that for early-stage patients, the baseline features of actual progressors are completely invisible to the algorithms, leading the model to confidently default to the majority negative class and driving the global evaluation’s failure.

**Figure 2:**
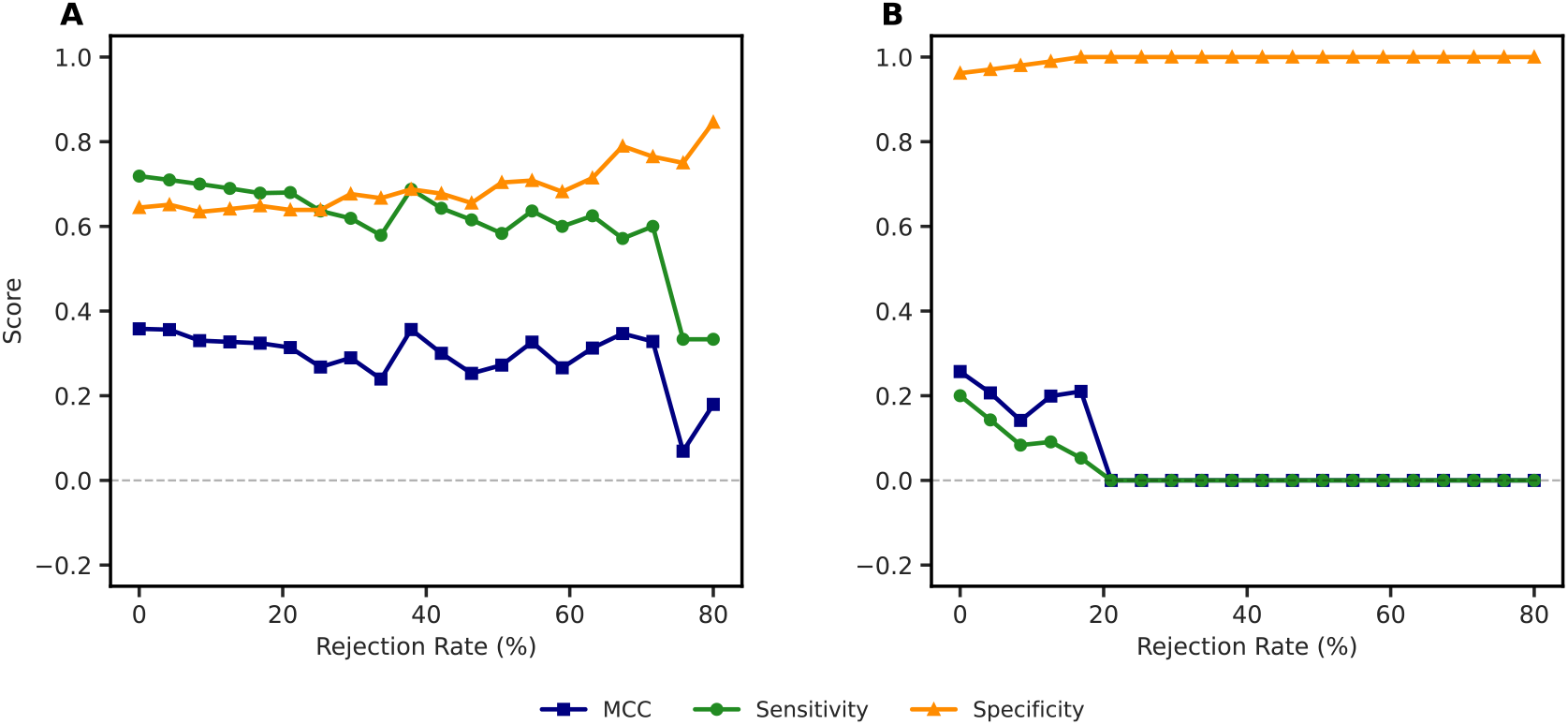
tratified rejection dynamics by discovered subgroup. Performance metrics evaluated independently across rejection thresholds. **A**. Subgroup A maintains stable, positive predictive performance even at 70% rejection, indicating a hard performance ceiling where advanced baseline symptoms provide limited but reliable discriminative signal. **B**. Subgroup B exhibits a catastrophic collapse: at ≥20% rejection, the ensemble completely ceases predicting the positive class (MCC = 0), leaving a pool of confident False Negatives. This identifies the early-stage clinical profile as the root cause of the dataset’s global discriminative failure.

## 4 Conclusion

This study shows that the apparent failure of uncertainty-informed rejection in LID forecasting is driven by a specific subgroup of early-stage patients. Stratified evaluation revealed that global performance metrics can be misleading, masking two distinct predictive regimes. In patients with moderate-to-advanced symptoms at baseline, uncertainty was partly resolvable and models could extract a reliable prognostic signal, though with a clear performance ceiling. In contrast, baseline features in mild, early-stage patients lacked sufficient discriminative power, making future trajectories effectively invisible to the models. As a result, predictions defaulted to the majority class, and true progressors were confidently misclassified as negative. These findings suggest that improving early-stage LID forecasting will require moving beyond static baseline clinical data toward longitudinal measures, wearable data, or molecular biomarkers.

## Data Availability

All datasets and code supporting the findings of this study are available from the authors upon reasonable request.

